# Comparative analysis of simplified and standard protocols for managing moderate and severe acute malnutrition in outpatient services in Venezuela: a prospective cohort study

**DOI:** 10.64898/2026.04.27.26351902

**Authors:** Pablo Hernandez, Claret Mata, Mary Lares, Melvin Morán, Zulay González, Elisabete Catarino, Yaliska Ramos, Alegna Varela, Yvette Fautsch-Macías, Grace Funnell

**Affiliations:** Department of Nutrition and Food Sciences, Nutrition and Dietetics School, Faculty of Medicine, Central University of Venezuela, Caracas, Capital District, Venezuela; Department of Public Health Sciences, Nutrition and Dietetics School, Faculty of Medicine, Central University of Venezuela, Caracas, Capital District, Venezuela; Health and Nutrition Section, The United Nations Children’s Fund, Venezuela. Caracas, Capital District, Venezuela; Survive and Thrive Section, The United Nations Children’s Fund, Regional Office for Latin America and the Caribbean, Panama City, Panama; Child Nutrition and Development Programme Group, The United Nations Children’s Fund - Headquarter. New York City, New York, USA

## Abstract

To provide adequate care to children with acute malnutrition, different management protocols have been in use, including the WHO standard guideline and a simplified protocol. The latter, used in Venezuela since 2020, has adopted three simplifications: 1) Expanded criteria for treatment admission; 2) Use of a single treatment product; 3) Simplified dosage: use of 2 sachets per day to treat SAM cases and 1 sachet per day to address MAM cases, regardless of weight. Our study compares the effectiveness, length of stay and programmatic costs of the simplified protocol and the WHO standard guideline in addressing acute malnutrition in children aged 6-59 months in Venezuela from February to August 2024. A total 229 children were enrolled in a prospective cohort study. Monitoring was continuous up to 16 weeks of treatment, evaluating key indicators including weight gain, recovery time, survival, recovery and default rates, number of sachets of RUTF consumed and implementation costs. Baseline characteristics were similar between cohorts, with most cases being moderate in both the standard (90.9%) and simplified (86.6%) cohorts. Both protocols demonstrated similar anthropometric improvements and recovery trajectories throughout follow-up, with no significant differences (p > 0.05). However, the simplified protocol showed higher recovery rates overall for MAM and SAM (70.1% vs 59.4%, p=0.031), although default rates remained high in both protocols (24.8% in the standard protocol vs. 18.7% in the simplified protocol). The simplified protocol presented reduced costs by 15% ($133 vs $157 per recovered child). These results suggest that the simplified protocol, using a single product and an adapted dosage, is as effective as the standard protocol for treating children with acute malnutrition in Venezuela. The findings support wider implementation of the simplified protocol particularly in resource-limited settings. Further research is needed to optimize protocols and improve adherence to reduce default rates.

## Introduction

Acute malnutrition, encompassing both moderate acute malnutrition (MAM) and severe acute malnutrition (SAM), remains a significant global health challenge, particularly in developing countries [1]. It affects millions of children under five years of age worldwide, with an estimated 52 million children suffering from acute malnutrition annually [2]. SAM and MAM have serious consequences, contributing to high rates of morbidity and mortality among children [3].

The prevalence and impact of acute malnutrition varies across regions. In 2022, global data showed that 14.8% of children under five in South Asia suffered from acute malnutrition. In West and Central Africa, the prevalence was 6.9%, while in Latin America, it reached 1.4% [4]. A national report, published by the National Nutrition Institute in 2009 indicates that 4.1% of children under five in Venezuela suffered from acute malnutrition [5]. This is last national figure, as the country has not released updated official reports since then.

The absence of comprehensive, nationally representative prevalence data in official statistics has necessitated reliance on alternative sources, such as programmatic data from non-governmental organizations. Despite utilizing WHO’s cut-off points for both weight for height and mid-upper arm circumference (MUAC), unofficial reports may not accurately reflect the national population prevalence due to potential biases inherent in data collected from local passive screening programs. For example, a previous report in 2021 [6], analyzed a non-representative sample of 46,462 children under five years, screened between 2017 and 2019, by Caritas Venezuela in the poorest parishes of 15 out of the 23 Venezuelan states. Caritas used the WHO 2006 child growth standards to identify acute malnutrition and found a decrease in acute malnutrition rates between 2017 and 2019 from 14.8% to 11.5% for MAM and from 4.9% to 3.4% for severe acute malnutrition.

On the other hand, Candela’s research [7] conducted between 2019 and 2020, assessed an intentional (non-probabilistic) sample of 1851 children under five years participating in community food and nutrition programs in the Capital District, Miranda, Carabobo, Lara, and La Guaira states. This study employed the standard anthropometric approach used by Caritas applying WHO cut-off points for both weight for height and mid-upper arm circumference (MUAC) to classify nutritional status. The study found a proportion of 3% of MAM and 2% of SAM among beneficiaries. The highest proportion of acute malnutrition was found in Carabobo (10%), Miranda (7%), the Capital District (6%), and La Guaira (6%).

The WHO standard protocol is based on recommendations from the WHO 2013 guideline [8]. In June 2023, the WHO updated the guideline for the prevention and management of acute malnutrition in children under 5 years of age [9], proposing that during care, the child should not be considered in isolation but taking into account its environment. In these new guidelines, the WHO recommends SAM cases to be treated with RUTF, and MAM cases to be evaluated holistically, factoring in medical and food security considerations to determine whether management with specially formulated foods (SFFs) is necessary, particularly in contexts where children exhibit a higher mortality risk. Notably, the WHO guidelines recognize the necessity of adapting protocols to better respond to diverse and unique environmental contexts.

In response to the COVID-19 pandemic in 2020, Venezuela implemented simplified adaptations to the 2013 WHO standard protocol, which was the official guideline at that time. These adjustments involved streamlining procedures for the diagnosis and treatment of acute malnutrition to ensure continuity of care during the health emergency, including: 1) Expanded criteria for treatment admission: increased mid-upper arm circumference (MUAC) cut-offs to admit all children <125 mm, and/or weight-for-height/length Z Score < -2 SD, and/or bilateral pitting oedema; unlike the standard protocol the simplified protocol one broadens treatment eligibility criteria by encompassing both SAM and MAM; 2) Use of a single treatment product: the simplified protocol used in Venezuela prescribes RUTF for all both MAM and SAM cases. This differs from the 2013 WHO standard guidelines, which limited RUTF to SAM cases, and the 2023 update, which now recommends Specialized Nutritional Foods (SNF) specifically for high-risk MAM cases; 3) Simplified dosage: the simplified protocol one prescribes 2 sachets per day for SAM cases and 1 sachet per day for MAM cases, regardless of weight, while the WHO standard protocol recommends to use a weight-based RUTF dosage [10].

To-date, no studies have investigated the effectiveness of the use of a simplified protocol for the treatment of acute malnutrition in Venezuela. The lack of scientific publications comparing both protocols for the treatment of acute malnutrition presents an opportunity to generate evidence and improve care for this condition in the country.

This study aims to compare the effectiveness, length of stay and programmatic costs between the simplified protocol, which includes the three adaptations cited above and the standard protocol based on the 2023 WHO guidelines for the prevention and treatment of acute malnutrition in children aged 6 to 59 months, in outpatient care services across the Venezuelan states of Bolívar, Capital District, La Guaira, and Miranda.

## Materials and methods

### Study setting

This study was conducted across urban slum areas of four states of Venezuela (Bolívar, Capital District, La Guaira, and Miranda) from February 19 to August 31, 2024. These states encompassed 23.6% of the total Venezuelan population. The states were selected intentionally based on operational considerations, including proximity to the capital city (Caracas/Capital District), as well as the highest number of acute malnutrition cases identified by the nutrition programme supported by UNICEF in the country. Miranda and La Guaira, located in the northern region near the capital, facilitated coordination of research activities and access to resources. Bolívar, in the southern part of the country, is characterized by having the largest number of identified cases of acute malnutrition. Conducting the study in regions with varying malnutrition levels enabled assessment of the interventions’ outcomes across diverse epidemiological contexts within Venezuela.

### Study design and participants

This study was an observational, prospective cohort investigation that compared the simplified protocol in use in Venezuela with the 2023 WHO standard protocol among children aged 6-59 months with SAM or MAM.

Critically, the investigators did not assign individual participants or healthcare centers to specific protocols. Instead, the protocol used at each site was determined prior to the study by the National Public Health Network, including the Bolivar State Public Health Institute, and the non-governmental organization Fe y Alegría, based on their internal operational guidelines and regional health policies.

The researchers served as independent evaluators of these pre-established programmatic implementations. Both protocols were executed concurrently across the four states, with RUTF serving as the sole therapeutic product for all participants.

A purposeful sampling was used. The sample comprised all children who attended the outpatient care centers of both - the national public health network, including the Bolivar State Public Health Institute, and the care centers of Fe y Alegría - a national non-governmental organization that implements and supports nutrition programs - who met the study’s inclusion criteria during the three-month data collection period for each cohort, whether identified directly at the care centers or during community screening activities.

To facilitate comparison between the simplified and the WHO standard protocols, Table 1 summarizes their main features, including eligibility criteria, treatment regimens, and recovery definition.

**Table 1.** Comparison between the simplified protocol and the standard protocol for the prevention and treatment of uncomplicated acute malnutrition in children under 5 years of age.

| Aspects | Simplified Protocol | WHO Standard Protocol |
| --- | --- | --- |
| Eligibility criteria | One of the following:<br>• MUAC <125 mm or<br>• Weight-for-Height/length Z Score < -2 SD or<br>• Bilateral pitting oedema<br>And all the following:<br>• No medical complications<br>• Positive appetite test | **SAM:**<br>• MUAC <115 mm or<br>• Weight-for-Height/length Z Score < -3 SD or<br>• Bilateral pitting oedema<br>And all the following:<br>• No medical complications<br>• Positive appetite test<br>**MAM:**<br>• MUAC ≥ 115 mm to < 125 mm or |
|  |  | • Weight-Height/length Z Score $\geq -3$ SD to $< -2$ SD and<br>• No nutritional oedema<br>And at least one the following:<br>• Failing to recover from moderate wasting after receiving other interventions (counselling alone).<br>• Having relapsed to moderate wasting<br>• History of severe wasting<br>• Households' low socioeconomic status (determined by the local Graffar-Mendez-Castellanos method [11])<br>• Co-morbidity (HIV, tuberculosis, or a physical or mental disability that does not jeopardize the anthropometric measurements) |
| Product and daily dosage | • SAM: Two sachets of 92g of RUTF/day (1000 kcal/day approx.).<br>• MAM: One sachet of 92g RUTF/day (500 kcal/day approx.). | Weight-based dosing<br>• SAM: 150-185 kcal/kg/day is offered using RUTF until the patient passes to MAM and then the dose is reduced to 100 -130 kcal/kg/day using RUTF until recovery.<br>• MAM: 100-130 kcal/kg/day is offered using RUTF until recovery. |
| Transition from SAM to MAM | When in one of the control measurements, the Weight-Height/length Z Score measurement is $\geq -3$ and $< -2$ and/or the MUAC is $\geq 115$ mm and $< 125$ mm, there is no oedema, the appetite is preserved, and the patient is clinically stable and alert. |  |
| Follow-up duration | Min: 4 weeks<br>Max: 16 weeks | Treatment is offered from admission until full recovery. However, A cut-off was considered after 16 weeks of treatment/enrollment to ensure comparison with the other group |
| Recovery criteria | • No bilateral pitting oedema<br>• Clinically stable and alert.<br>One of the following two consecutive visits:<br>• If the child was admitted with MUAC < 125 mm, to leave he must have a MUAC ≥ 125 mm<br>• If the child was admitted with a Weight-Height/Length Z Score < -2, to exit the protocol they must have a Weight-Height/Length Z Score ≥ -2 | • No bilateral pitting oedema<br>• Clinically stable and alert.<br>All the following two consecutive visits:<br>• MUAC ≥ 125 mm<br>• Weight-height/length Z Score ≥ -2 |
| Default | Absent for consecutively for 2 follow-up visits. |  |
| Non-recovered | When the child does not reach the recovery criteria for the cut made after 16 weeks of treatment. [In both cohorts, a cut-off at 16 weeks of treatment is contemplated to ensure comparison between groups.] |  |
| Exit procedures | An exit dose of 7 sachets of RUTF is offered. | Nutritional counseling is offered. |

Children, with generalized oedema, who did not pass the appetite test, or with any other medical complications requiring specialized inpatient care, those who planned to move out of the area in the next 4 months and those with known allergies to any of the components of the RUTF were excluded from the study.

### Procedures - study outcomes

The study involved two cohorts of children aged 6–59 months diagnosed with acute malnutrition: one treated using the simplified protocol used in Venezuela and the other following the WHO standard protocol. Both cohorts were managed concurrently across the four study sites.

In both cohorts, trained health personnel conducted daily screenings of children attending care centers or participating in community screening sessions. After informed consent was sought from the parents or caregivers, pertinent data for the research were collected using a digital data collection form.

The initial evaluation of both cohorts involved collection of demographic (children’s sex, age, and ethnicity; parent or caregiver sex and age), socioeconomic (educational level of the mother, households’ socioeconomic status (SES), and households’ size), anthropometric (weight, length for children under 2 years old or height for children of two or more years, and MUAC measured according with to WHO and UNICEF standardized methodology [12], as well as clinical data (medical history, appetite and presence of oedema), and morbidity (adverse events, including fever, cough, diarrhea, emesis or allergic reactions). Subsequently, the nutritional intervention was administered based on the protocol criteria (see Table 1). Children who visited public health outpatient clinics were administered the simplified protocol, while those at Fe y Alegria care centers received the standard protocol.

Follow/up visits were conducted biweekly for children with MAM and weekly for those with SAM for children in both cohorts. During these appointments, anthropometry and clinical observations throughout the treatment period were recorded. Adherence to the prescribed regimen was requested, and a subsequent quantity of RUTF was provided to last until the next follow-up scheduled visit. The primary outcome of the study was nutritional recovery, and the secondary outcomes included default, non-recovery, referral to inpatient care, and mortality (see definitions in Table 1).

Upon recovery, children were medically discharged from the program. The exit procedures are described in Table 1.

### Effectiveness assessment

The effectiveness of both protocols—defined as their ability to improve nutritional status and promote appropriate growth in children with acute malnutrition was evaluated using the weight gain indicator, operationalized as weight change (g per kg per day) from treatment initiation to recovery.

Additionally, as a secondary outcome we included active surveillance of potential side events throughout the treatment period. This involved monitoring for gastrointestinal disturbances such as diarrhea, emesis, abdominal pain, and allergic reactions, including rash, urticaria, or pruritus. A clinical examination was conducted at each visit.

### Length of stay assessment

Length of stay, was defined as the number of weeks from treatment initiation to recovery.

### Programmatic cost assessment

Programmatic cost data were collected from a societal perspective, drawing on available accounting records and a series of interviews with key informants -including health workers, civil society organizations, parents or caregivers, and UNICEF staff- to capture all relevant costs incurred by institutions, beneficiaries and communities.

Only costs directly related to MAM and SAM treatment during the six-month study period were considered. Research-related costs (e.g., research team salaries, supervision, training and materials) were excluded.

Personnel costs included only health workers directly attending children, weighted by the number of centers and staff involved in each protocol. Program costs included office materials, RUTF, anthropometric equipment (height boards, weight scales and MUAC tapes) and costs of training and field supervision of the activities. Community contributions included transportation and food expenses incurred on consultation days as well as caregiver’s loss of productivity (or opportunity cost), estimated as potential income lost due to participating in follow-up visits. Total costs were aggregated and presented in 2024 US$ per treated and recovered child.

### Quality control and supervision

Quality control of the measurements and data collection was ensured by local supervisors who accompanied the measurement and treatment procedures at the healthcare centers, verifying the correct diagnoses and management of supplies throughout the collection through a rotation process programmed by the assigned centers. Regional supervisors ensured the accuracy of the data; they identified and corrected errors and inconsistencies, and conducted regular field visits, both scheduled and unscheduled. The research team continuously supervised data collection activities and was responsible for analyzing and reporting the findings. A data manager reviewed the databases resulting from real-time inconsistency identification and correction process to conduct a final quality assurance exercise.

### Ethics

The research protocol was approved by the independent bioethics committee of the National Bioethics Center of Venezuela (CIBI-CENABI #10/2023, February 16th, 2024). Data collection and nutritional care procedures adhered to the international bioethical considerations of the Declaration of Helsinki and Code of Ethics for Life of the National Fund for Science, Technology and Innovation (FONACIT) of the Bolivarian Republic of Venezuela. The trial was registered at ClinicalTrials.gov (NCT06287827). Written informed consent was obtained from parents and caregivers at the time of children’s admission to the study.

### Statistical analysis

Data were stored in an electronic database and analyzed using the Statistical Package for Social Sciences (SPSS) version 26. Descriptive summaries of participant characteristics and outcomes were presented using means and their standard deviations (SD), medians and interquartile ranges (IQR) for non-normally distributed continuous variables, and frequencies (n) and percentages for categorical variables. The Mann-Whitney U test was used to estimate the statistical significance of non-normally distributed continuous variables between protocols. Categorical variables were described in terms of frequency and compared between protocols using the χ2 test or Fisher’s exact test. Kaplan-Meier survival curves and log-rank tests were used to predict the time to recovery and compare survival curves across the protocols. Bivariable and multivariable Cox proportional hazard regression models were fitted to identify independent predictors of nutritional recovery. Multicollinearity was assessed prior to regression analysis. Variables for inclusion in the multivariate Cox proportional hazards model were selected through bivariate analysis, using a p-value cutoff of ≤ 0.25. Predictors of recovery were identified as statistically significant in the final multivariable model based on a p-value threshold of 0.05.

## Results

A total of 4,649 children were screened; of these, 7.3% were identified with Acute Malnutrition (n = 339 children), of whom 87,6% had MAM (145 boys and 152 girls) and 12,4% had SAM (22 boys and 20 girls). A total of 40 children were not eligible for the study: 8 of them had a negative appetite test, 11 presented a pre-existing medical condition (e.g., hypercalciuria, congenital heart disease, generalized edema), 16 caregivers/representatives did not sign the informed consent, and 5 resided outside the screening areas, representing a total of 11.8% of those identified. For this reason, 299 children (6.4% of total screened) were admitted to the study (Fig 1).

**Fig 1.**
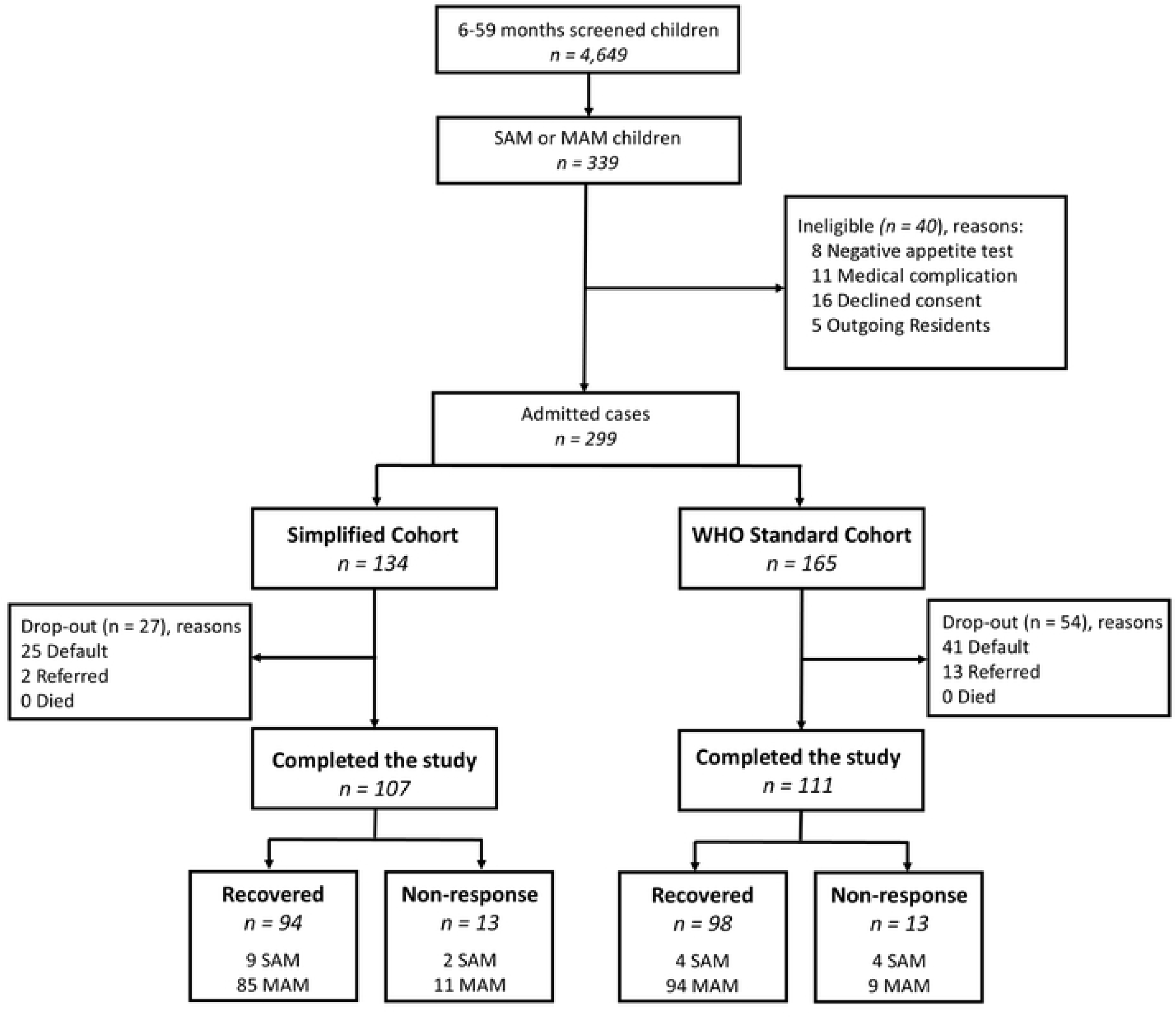
Flowchart of children admitted and treated under each study protocol. WHO, World Health Organization; SAM, severe acute malnutrition; MAM, moderate acute malnutrition.

### Baseline characteristics

Table 2 presents the baseline characteristics of children and their households in both protocols at study admission. The two groups were comparable, exhibiting similarities across most baseline variables, including demographic factors, anthropometric indicators, and acute illness symptoms. The only statistically significant differences were observed in maternal age, with mothers in the Standard protocol group being slightly older (p = 0.042). Additionally, a smaller proportion of children in the Simplified Protocol group had MUAC < 11.5 cm (p = 0.003), and a higher proportion presented oedema (p = 0.012). However, these significant differences are unlikely to have substantially influenced the study outcomes.

**Table 2.** Baseline characteristics of children admitted in the study.

| Characteristics | Standard protocol<br>(n = 165) | Simplified protocol<br>(n = 134) | *P* |
| --- | --- | --- | --- |
| Males, n (%) | 84 (50.9) | 64 (47.8) | 0.588 |
| Age in months, median (IQR) | 30.7 (15; 47) | 29.7 (15; 43) | 0.618 |
| Mother age, median (IQR) | 29 (24; 36) | 27.0 (22.8; 34) | 0.042 |
| Mother is caretaker, n (%) | 138 (83.6) | 121 (90.3) | 0.092 |
| Low SES, n (%) | 135 (81.9) | 111 (82.8) | 0.118 |
| History of hospitalization, n (%) | 5 (3.0) | 4 (3.0) | 0.982 |
| History of severe wasting, n (%) | 28 (17.0) | 11 (8.2) | 0.025 |
| Household size, median (IQR) | 5 (4; 6) | 5 (4; 6) | 0.981 |
| Weight (kg), median (IQR) | 9.3 (7.4; 12.3) | 9.3 (7.9; 11.8) | 0.797 |
| Height/length (cm), median (IQR) | 84.1 (73.3; 98.5) | 83.3 (74.8; 95.5) | 0.768 |
| WHZ/WLZ Score, median (IQR) | -2.18 (-2.52; -2.07) | -2.17 (-2.39; -2.05) | 0.420 |
| WHZ/WLZ < -3 SD, n (%) | 15 (9.1) | 9 (6.7) | 0.452 |
| WHZ/WLZ < -2 SD, n (%) | 128 (77.6) | 110 (82.1) | 0.335 |
| MUAC (cm), median (IQR) | 13.6 (12.4; 14.1) | 13.4 (12.5; 14.0) | 0.421 |
| MUAC 11.5 to <12.5 cm, n (%) | 2 (1.2) | 6 (4.5) | 0.081 |
| MUAC < 11.5 cm, n (%) | 40 (24.2) | 15 (11.2) | 0.003 |
| Oedema, n (%) | 0 (0.0) | 5 (3.7) | 0.012 |
| HAZ Score, median (IQR) | -1.41 (-2.58; -0.68) | -1.41 (-2.34; -0.80) | 0.942 |
| HAZ < -3 SD, n (%) | 29 (17.6) | 19 (14.2) | 0.426 |
| HAZ < -2 SD, n (%) | 31 (18.8) | 27 (20.1) | 0.767 |
| Severe acute malnutrition, n (%) | 15 (9.1) | 18 (13.4) | 0.233 |
| Mother reports fever, n (%) | 24 (14.5) | 28 (20.9) | 0.150 |
| Mother reports cough, n (%) | 83 (50.3) | 60 (44.8) | 0.341 |
| Mother reports diarrhea, n (%) | 18 (10.9) | 9 (6.7) | 0.208 |
| Mother reports emesis, n (%) | 6 (3.6) | 3 (2.2) | 0.482 |

Abbreviations: IQR, Interquartile Range; SES, Socioeconomic Status; WHZ, Weight-for-Height Z-score; WLZ, Weight-for-Length Z-score; MUAC, Mid-Upper Arm Circumference; HAZ, Height-for-Age Z-score.

### Treatment outcomes

Table 3 shows the treatment outcomes for children included in the study, categorized by the protocol (Standard or Simplified) and by the severity of malnutrition (total, SAM and MAM). The Simplified Protocol had a significantly higher recovery rate (70.1% overall and 73.3% among MAM cases) compared to the Standard Protocol (59.4% and 62.7% respectively). No statistically significant differences were observed among the SAM cases. The Simplified Protocol had significantly fewer referrals to inpatient care.

**Table 3.** Study outcome indicators in total, SAM and MAM children by protocol followed.

| **Outcome** | **Standard protocol**<br>**(n = 165)** | **Simplified**<br>**protocol**<br>**(n = 134)** | ***P*** |
| --- | --- | --- | --- |
| **All children** |  |  | **0.031** |
| Recovered | 98 (59.4) | 94 (70.1) |  |
| Non-response | 13 (7.9) | 13 (9.7) |  |
| Default | 41 (24.8) | 25 (18.7) |  |
| Referred to inpatient<br>care | 13 (7.9) | 2 (1.5) |  |
| **Children with SAM** |  |  | **0.177** |
| Recovered | 4 (26.7) | 9 (50.0) |  |
| Non-response | 4 (26.7) | 2 (11.1) |  |
| Default | 2 (13.3) | 5 (27.8) |  |
| Referred to inpatient<br>care | 5 (33.3) | 2 (11.1) |  |
| **Children with MAM** |  |  | **0.014** |
| Recovered | 94 (62.7) | 85 (73.3) |  |
| Non-response | 9 (6.0) | 11 (9.5) |  |
| Default | 39 (26.0) | 20 (17.2) |  |
| Referred to inpatient<br>care | 8 (5.3) | 0 (0.0) |  |

Table 4 presents the changes in anthropometric measurements (weight, height, and MUAC) among children in both protocols, categorized by recovery status. In general, both groups showed similar gains and gain velocities in weight and MUAC with no significant differences between protocols. However, when looking at recovery, the Simplified protocol group had a significantly higher rate of height/length gain compared to the Standard Protocol group (<0.001)

**Table 4.**
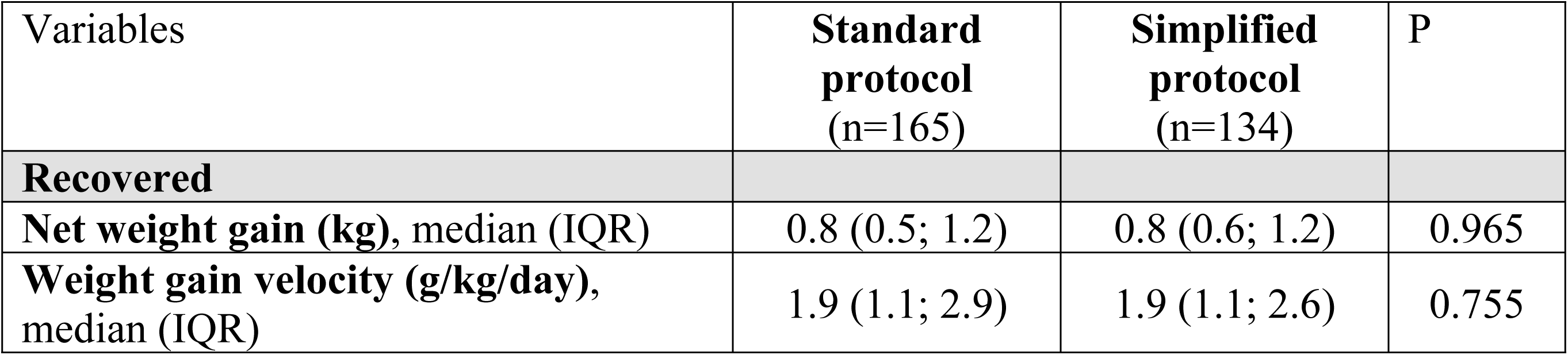
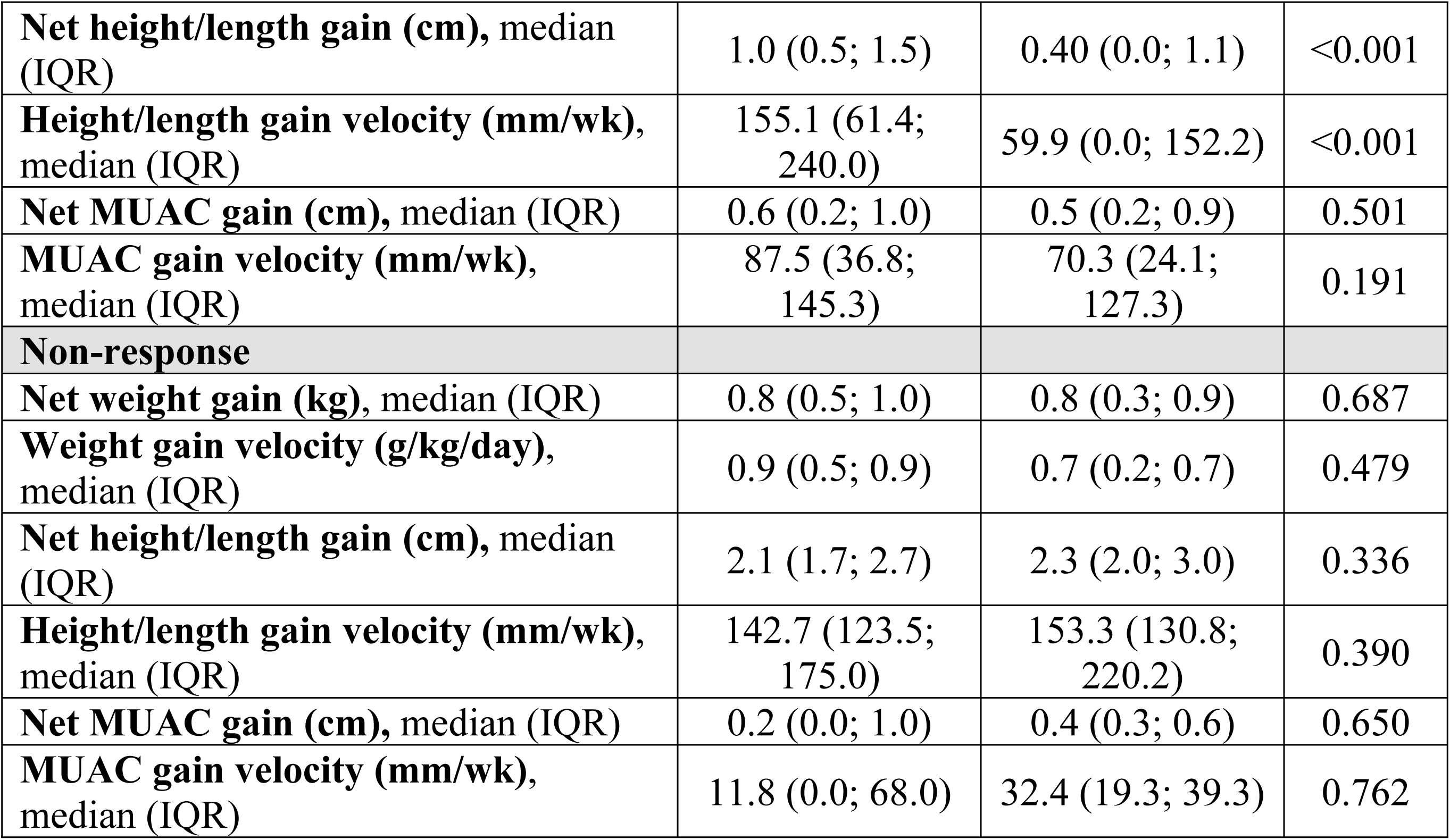
Comparison of changes in Anthropometric Measurements in recovered and non-response children by protocol followed.

### Recovery time from acute malnutrition and the Kaplan-Meier survival curve

The cumulative percentage of recovered cases per week of treatment is shown in Fig 2 using a Kaplan-Meier curve, illustrating nutritional recovery as the primary outcome. Recovery trajectories were comparable between protocols. By week 4, approximately 20% of children attained recovered status – mainly those with MAM – under both the Standard and Simplified protocols. Subsequently, both curves remained nearly parallel, indicating similar recovery rates between protocols (p= 0.982) throughout treatment considering the length of stay.

**Fig 2.**
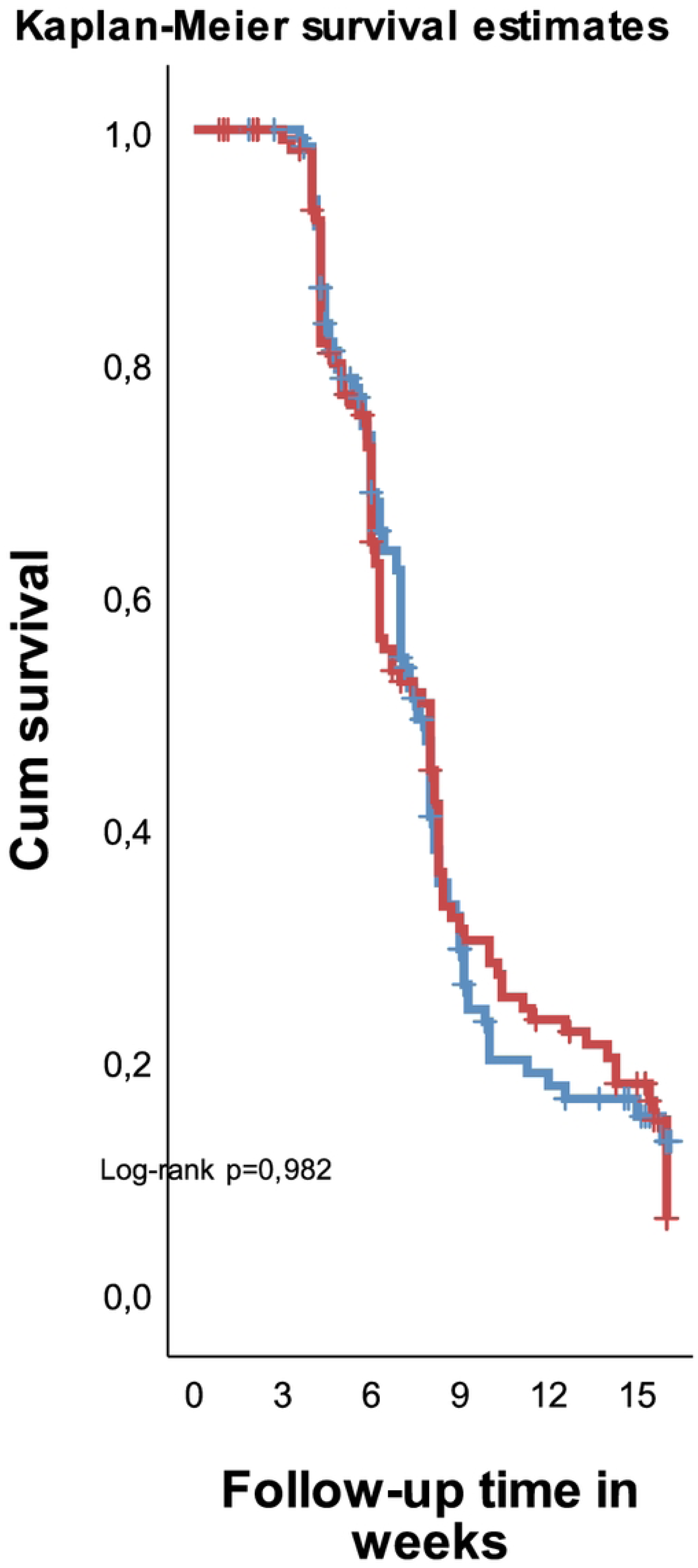
Kaplan-Meier survival curves comparing recovery time between protocols.

Table 5 provides information on the length of stay and the number of visits for children in the Standard and Simplified protocols. The median length of stay was similar across protocols, reinforcing the previous finding of no significant difference in recovery time. While the study primarily focused on outpatient care, a subset of participants required referral to inpatient facilities due to lack of clinical progress or the presence of any exclusion criteria; these cases exhibited significantly longer periods of outpatient management under the Standard protocol before referral (p = 0,019). The Simplified Protocol had a higher proportion of children with >4 visits (p = 0,019) and a higher percentage of on-time visits among the recovered children (p = 0,033).

**Table 5.** Length of stay and number of visits by protocol followed.

| Variables | Standard protocol<br>(n=165) | Simplified protocol<br>(n=134) | P |
| --- | --- | --- | --- |
| **Length of stay (wk), median (IQR)** |  |  |  |
| Recovered | 7 (5; 8) | 6 (5; 8) | 0,590 |
| Non-response | 15 (15; 16) | 15 (15; 16) | 0,418 |
| Default | 2 (2; 6) | 2 (2; 5) | 0,267 |
| Referred to inpatient care | 4 (2; 5) | 1 (1; 1) | 0,019 |
| **Number of visits, n (%)** |  |  | 0,019 |
| None | 24 (14,5) | 11 (8,2) |  |
| 1 – 2 | 92 (55,8) | 65 (48,5) |  |
| 3 – 4 | 34 (20,6) | 31 (23,1) |  |
| $> 4$ | 15 (9,1) | 27 (20,1) | |
| **Percentage of visit on time, median (IQR)** |  |  |  |
| Recovered | 50 (0; 100) | 75 (50; 100) | 0,033 |
| Non-response | 55,5 (40; 66,6) | 71,4 (60; 83,3) | 0,336 |
| Default | 33,3 (0,0; 75,0) | 66,6 (50; 100) | 0,049 |
| Referred to inpatient care | 50,0 (0,0; 70,8) | 100 (100; 100) | 0,088 |

Fig 3 demonstrates a statistically significant difference in the quantity of RUTF envelopes distributed between standard and simplified protocols across all treatment outcomes.

**Fig 3.**
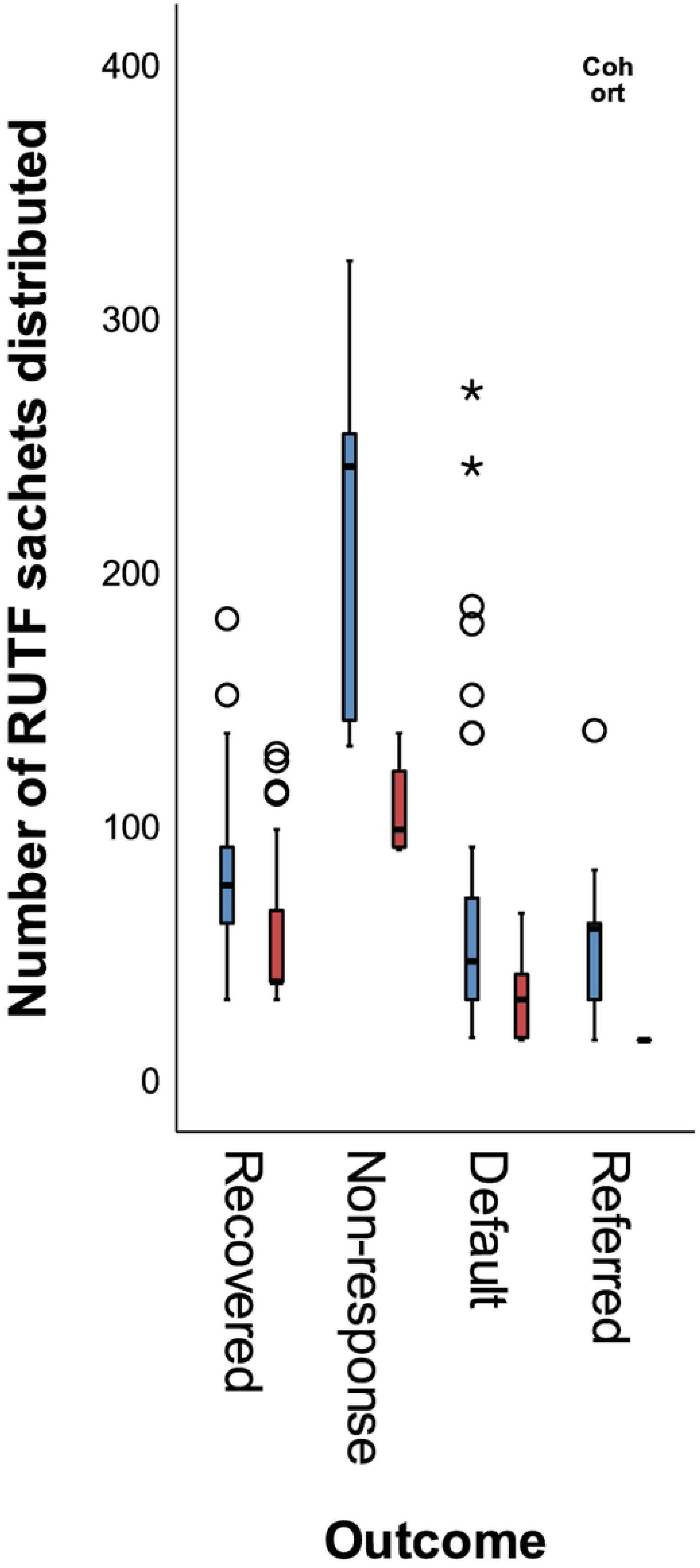
Number of RUTF sachets distributed to admitted cases according to the treatment outcome per protocol followed

Recovered children in the Standard Protocol received a median of 75 RUTF sachets (75; IQR: 60-90) compared with 37 sachets in the Simplified protocol (37; IQR: 37-65). This disparity was further accentuated in the non-recovered children: 240 RUTF sachets (IQR: 140-253), versus 97 sachets (IQR: 90-120) , representing a reduction of over 50% in the Simplified Protocol.

### Recovery predictors: Cox regression analysis

A Cox proportional hazards regression model was employed to identify factors associated with recovery from acute malnutrition. Variables exhibiting a p-value < 0.25 in bivariate analyses were included in the multivariable model. The final adjusted model encompassed socioeconomic status, number of visits, percentage of on-time visits, number of RUTF sachets distributed, baseline nutritional status, and admission criteria (Table 6).

**Table 6.** Predictors of recovery among children with severe and moderate acute malnutrition (Bi-variable and multi-variable cox regression analysis).

| Variables | Crude HR (95% CI) | Adjusted HR (95% CI) |
| --- | --- | --- |
| **Cohort** |  |  |
| Standard | Ref | - |
| Simplified | 0.997 (0.750, 1.324) | - |
| **Sex** |  |  |
| Male | Ref | - |
| Female | 1.103 (0.831, 1.465) | - |
| **Age in months** |  |  |
| 6–11 | Ref | - |
| 12–23 | 1.223 (0.741, 2.020) | - |
| 24–35 | 0.995 (0.575, 1.720) | - |
| 36–59 | 0.986 (0.600, 1.620) | - |
| **SES** |  |  |
| Strata II | Ref | Ref |
| Strata III | 1.258 (0.476, 3.327) | 66.708 (19.160, 232.254) |
| Strata IV | 1.762 (0.719, 4.315) | 64.671 (20.056, 208.527) |
| Strata V | 1.372 (0.543, 3.470) | 85.536 (25.550, 286.360) |
| **Number of visits** | 0.427 (0.364, 0.500) | 0.149 (0.113, 0.198) |
| **Percentage of visit on time** | 1.006 (1.002, 1.010) | 1.037 (1.031, 1.043) |
| **Number of RUTF sachets distributed** | 0.975 (0.970, 0.980) | 0.990 (0.985, 0.996) |
| **Baseline nutritional status** |  |  |
| MAM | Ref | Ref |
| SAM | 0.475 (0.270, 0.834) | 2.185 (1.109, 4.303) |
| **Admission criteria** |  |  |
| WHZ | Ref | Ref |
| MUAC | 0.798 (0.544, 1.168) | 1.326 (0.891, 1.973) |
| Edema | 7.399 (2.330, 23.499) | 5.712 (1.482, 22.007) |

After adjusting for all other variables in the model, the children from households in the lowest SES stratum (V) exhibited a nearly 86-fold increased hazard of recovery compared to those in the highest stratum (II), this estimate was accompanied by substantial uncertainty, as reflected in the wide 95% confidence interval (25.550, 286.360).

In the adjusted model, an increased number of visits was significantly associated with the recovery outcome. Each additional visit correlated with an approximate 85% reduction in the hazard of recovery. Conversely, a 10% increase in the percentage of on-time visits was associated with a 37% increase in the hazard of recovery. Each additional RUTF sachet distributed was associated with a 1% reduction in the hazard of recovery. Children presenting SAM at baseline exhibited a more than twofold increased hazard of recovery compared to those with MAM.

Regarding admission criteria, children admitted based on the presence of edema demonstrated a statistically significant increased hazard of recovery compared to those admitted based on WHZ criteria. No statistically significant difference in recovery hazard was observed between children admitted based on MUAC criteria and those admitted based on WHZ.

The presence of side events was monitored at each visit throughout the treatment period, up to a maximum 16 weeks. Cough was the most frequently reported symptom (63,4% in Simplified cohort versus 64,8% in Standard cohort), followed by fever (41,8% in simplified cohort versus 30,9% in standard cohort) and diarrhea (23,1% in simplified cohort versus 25,5% in standard cohort), with no statistically significant differences between cohorts (Table 7). Allergic reactions were observed in 3% of participants in the Simplified protocol and 6% in the Standard protocol. All reactions were transient, resolved without sequelae, and, upon investigation by the medical team, were determined to be unrelated to the RUTF or any of its constituent ingredients.

**Table 7.** Presence of side events along the treatment period of children by protocol followed.

| Side events | Standard protocol<br>(n=165) | Simplified protocol<br>(n=134) | p |
| --- | --- | --- | --- |
| **Cough** |  |  | 0.800 |
| Yes | 107 (64.8) | 85 (63.4) |  |
| No | 58 (35.2) | 49 (36.6) |  |
| **Fever** |  |  | 0.051 |
| Yes | 51 (30.9) | 56 (41.8) |  |
| No | 114 (69.1) | 78 (58.2) |  |
| **Diarrhea** |  |  | 0.642 |
| Yes | 42 (25.5) | 31 (23.1) |  |
| No | 123 (74.5) | 103 (76.9) |  |
| **Vomit** |  |  | 0.204 |
| Yes | 34 (20.6) | 20 (14.9) |  |
| No | 131 (79.4) | 114 (85.1) |  |
| **Allergic reactions** |  |  | 0.211 |
| Yes | 10 (6.1) | 4 (3.0) |  |
| No | 155 (93.9) | 130 (97.0) |  |

### Programmatic cost analysis

Table 8 details the estimated programmatic costs per treated and recovered child for each protocol. The Simplified protocol cost USD 109.48 per enrolled child and USD 133.04 per recovered child, compared with USD 122.25 per enrolled child and USD 157.31 per recovered child, respectively, for the Standard Protocol. This represents a cost differential of USD 12.77 per enrolled child and USD 24.27 per recovered child, with the Standard Protocol being more expensive. This increased cost in the Standard Protocol is primarily attributable to higher program costs, particularly the greater number of RUTF used. Program costs accounted for the largest proportion of individual expenditures in both protocols. The most significant difference between the two was observed in RUTF-related expenses. The Standard Protocol, which used a greater number of RUTF sachets per child, resulted in RUTF treatment costs approximately 1.5 times higher than those associated with the Simplified Protocol.

**Table 8.** Estimated programmatic cost per child treated and recovered according to protocol followed.

|  | Simplified |  |  |  | Standard |  |  |  |
| --- | --- | --- | --- | --- | --- | --- | --- | --- |
|  | *Treated* |  | *Recovered* |  | *Treated* |  | *Recovered* |  |
|  | USD | % of total costs | USD | % of total costs | USD | % of total costs | USD | % of total costs |
| **Personnel cost** | 12.38 | 11.31 | 12.76 | 9.59 | 11.66 | 9.53 | 11.95 | 7.59 |
| **Programme costs** | 76.68 | 70.04 | 97.43 | 73.23 | 93.46 | 76.45 | 125.17 | 79.57 |
| **Community contributions** | 20.42 | 18.65 | 22.85 | 17.17 | 17.13 | 14.01 | 20.19 | 12.84 |
| **TOTAL (US \$)** | 109.48 | 100.00 | 133.04 | 100.00 | 122.25 | 100.00 | 157.31 | 100.00 |

## Discussion

This prospective observational cohort study represents, to our knowledge, the first Latin American investigation comparing, in terms of efficacy, length of stay and programme cost, a simplified protocol with the WHO standard protocol for the management of acute malnutrition in exceptional circumstances, applying the most recent updated recommendations [9]. This absence of prior regional data highlights a critical gap in our understanding of how these protocols perform within the specific demographic and healthcare contexts of Latin America.

The cohorts evaluated under both protocols exhibited no statistically significant difference in demographic factors, anthropometric variables, or prior medical history. This natural homogeneity across key baseline characteristics strengthens the comparative power of our study, allowing us to attribute observed differences in outcomes more confidently to the distinct protocols themselves.

Our findings revealed a higher recovery rate with the simplified protocol, particularly in MAM cases, compared to the standard protocol. The results of the simplified protocol suggest potential mechanisms beyond mere caloric sufficiency, such as enhanced adherence to the care protocol or more effective integration into the national community-based care systems. Further exploration of these underlying factors is crucial to refine our understanding of optimal nutritional interventions according to context. Moreover, these results contrast with the conventional assumption that higher RUTF doses inherently translate to superior recovery outcomes for SAM [13], highlighting the need to consider other context-specific adaptations to the Standard Protocol for exceptional circumstances such as emergency and resource constrained contexts.

The higher recovery rate observed with the simplified protocol (70%) compared to the standard protocol (59%) in this study aligns with findings from other research conducted in certain regions of Africa, which face similar challenges, such as food insecurity, logistical barriers to healthcare access, and limited resources for nutritional interventions. Specifically, the ComPAS (Combined Protocol for Acute Malnutrition Study) [14], a single-blinded, randomized, controlled, non-inferiority trial in Kenya and South Sudan, demonstrated comparable recovery rates between a simplified combined protocol and the standard protocol (76.3% and 73.5%, respectively) in children aged 6-59 months, mirroring this study’s focus on both severe and moderate acute malnutrition and using the same product (RUTF). Furthermore, the Optima study [15], a non-inferiority, randomized controlled trial in the Democratic Republic of the Congo, revealed the superiority of a simplified strategy over the standard protocol, with 92% of children showing improvement at 16 weeks compared with 75% in the standard protocol (using separate protocols and products for severe acute malnutrition and moderate acute malnutrition). Additionally, cohort studies evaluating the simplified protocol alone in Mali [16], Niger [17], and Burkina Faso [18] reported recovery rates of 92.3%, 82.3%, and 84%, respectively. These diverse findings, spanning randomized controlled trials and observational cohorts in various tropical settings, collectively suggest that protocols using simplified approaches can achieve comparable, and in some cases, superior, recovery rates compared to standard protocols, particularly in resource-constrained environments.

It is important to recognize that the recovery rates observed in this study, although indicating a relative advantage for the simplified protocol (70% vs. 59%), did not meet the 75% threshold established by the Sphere Manual Project [19] for effective acute malnutrition management programs. This discrepancy is primarily due to the high percentage of defaulters, particularly within the standard protocol (24.8% vs. 18.7%). Similar challenges have been documented in other studies, such as the MANGO trial [20], which, despite focusing solely on severe acute malnutrition (SAM) and comparing reduced versus standard doses of RUTF, reported recovery rates below Sphere standards (52.7% and 55.4%, respectively). This was attributed to stringent hospital referral criteria, which accounted for nearly 20% of referral cases in the MANGO trial. Furthermore, a retrospective study in Ethiopia [21] among children with SAM revealed a recovery rate of 70.6%, with a high default rate of 17.84% cited as the primary contributing factor. This study also consistently exceeded the Sphere standard limit for default rates (<15%) with 24.8% in the standard protocol and 18.7% in the simplified protocol. These findings underscore the persistent challenge of patient retention in acute malnutrition programs, highlighting the need for targeted interventions to address factors contributing to treatment discontinuation, such as logistical barriers, social determinants of health, acceptability of the product, and the perceived burden of treatment [22], particularly in the resource-limited settings of Latin America. Other studies attributed the low default rate to the use of home visits [15],

Regarding efficacy, the study observed comparable weight gain velocity. Previous research, such as Bailey et al. [14], documented weight gains of 1.9 g/kg/day in both the standard and combined protocols, the same value that was found in this study both standard and simplified protocols.

The most common symptoms observed were cough, fever, and diarrhea in both cohorts, with no significant differences between them. These symptoms were transient, resolved without sequelae, and, upon investigation by the medical team, were determined to be unrelated to the RUTF or any of its constituent ingredients. Previous studies have indicated that diarrhea and vomiting are prevalent adverse effects in these programs, attributed to the ingestion of RUTF, whose lipid composition may lead to gastrointestinal disorders[23, 24]. Additionally, changes in feeding habits or inadequate hygiene practices in children may contribute to these effects[23, 25]. Furthermore, there were no reported fatalities among the children, likely because most cases were classified as MAM without serious criteria, and any complications were promptly referred to care centers.

In terms of efficiency, the duration of treatment was comparable across both protocols. A recovery peak was observed between weeks 4 and 8, corresponding to the recovery of most MAM cases, which typically required two to three consecutive follow-ups. Cazes et al. [15] reported that the mean time to achieve nutritional improvement was 8 weeks (RIC 5-12) in the standard group and 5 weeks (RIC 3-8) in the OptiMA group (p<0.001). Thus, the period between 4 and 8 weeks appears to be critical for recovery. The frequency of visits to the care center emerged as a significant factor in the children’s recovery. The simplified protocol had a higher proportion of children attending more frequently and punctually compared to the standard protocol. This may be due to the fact that, in the standard protocol, if more sachets per day were provided and not consumed within the designated time, caregivers might have opted to delay visits to the health center until the sachets were finished. The adjusted model indicated that a 10% increase in the percentage of timely visits enhances the likelihood of recovery by 37%. Furthermore, it was noted that extending a visit within the children’s care program reduces the chances of recovery by 85%. A previous study established that missing one visit predicted an extension of treatment duration by 3 weeks [16].

Finally, the programmatic cost of treatment using the simplified protocol was 15% lower than that of the standard protocol ($133.04 vs. $157.31 per recovery case). Beyond the individual benefits for children by enhancing the likelihood of positive outcomes, the simplified protocol appears to offer substantial advantages from a programmatic perspective as it facilitates the treatment of a greater number of children at a lower cost.

This study utilized routinely collected programmatic data, with a rigorous data tracking system implemented throughout to ensure the inclusion and follow-up of all children registered at health centers until their recovery. A key strength of this research lies in its demonstration of the feasibility of robust data tracking in programmatic context of four different states in Venezuela, which facilitates the creation of a high-quality database. The study design employed was longitudinal, which enabled the prospective observation and sequential data collection from the same cohort of children across the duration of the investigation. This methodological approach was essential for quantifying the changes achieved relative to the initial measurements recorded at the baseline assessment. Although selection bias could be present, the homogeneity in baseline characteristics across both groups ensured comparability, strengthening the validity of the results.

The limitations of this study must be also acknowledged. Although cohort studies offer valuable insights, they cannot establish definitive causality. One significant limitation is the reliance on self-reported secondary events, which may introduce recall and social desirability biases. This could potentially have affected the accuracy of the collected data. Another limitation is the lack of data on possible RUTF sharing among the participants and their relatives, which could have affected actual product consumption and outcomes. Additionally, the absence of data on the sensorial perception of the products limited our ability to assess the influence of palatability on adherence. Furthermore, post-treatment t follow-up data, including relapse and mortality, were not collected, which would have been valuable for assessing long-term effectiveness and identifying potential areas for improvement in post-treatment care.

## Conclusions

The results of our study suggest that the Simplified protocol may be as effective as the Standard protocol in addressing acute malnutrition, particularly among children with MAM in resource-constrained settings. Although both protocols demonstrated comparable recovery times, the Simplified protocol achieved equivalent weight gain with a reduced quantity of RUTF, leading to lower treatment costs per recovered case. Moreover, it showed higher recovery rates and fewer referrals to inpatient care. The Simplified Protocol may therefore offer advantages in treatment adherence and programmatic cost, potentially improving outcomes, especially for children with MAM. These results support the consideration of the Simplified protocol as a viable approach and alternative to optimize the coverage and cost-effectiveness of treatment in specific contexts. However, further research is necessary to validate these findings and explore the underlying mechanisms responsible for the observed differences, particularly in relation to treatment default and long-term outcomes.

## Acknowledgments

We express our gratitude to the children and their parents or caregivers who took part in this research. We sincerely thank the health staff members of Ministry of Popular Power for Health (MPPS) of Venezuela, the Bolivar State Public Health Institute, and the Fe y Alegría organization for their support in the implementation of this study. Also, we would like to extend our thanks to the healthcare workers and supervisors that provided treatment to study participants.

## Author contributions

Conceptualization: PH, CM, ML, MM, ZG, YR, AV; Methodology: PH, CM, ML, MM, ZG, YR, AV; Funding Acquisition: PH; Resources: MM, ZG, EC, YR, AV, GF; Project administration: CM; Investigation: PH, CM, ML; Supervision: ML; Data Curation: PH; Formal Analysis: PH; Visualization: CM; Validation: MM, ZG, EC, YR, AV, GF; Writing – original draft: PH, CM, ML, MM, ZG, EC, YR, AV, YF, GF; Writing – review & editing: PH, CM, ML, MM, ZG, EC, YR, AV, YF, GF.

## Data availability

All relevant data are presented within this paper. However, the raw data supporting this article is not publicly available, as participants did not provide explicit consent for data sharing. The dataset is securely stored in an encoded format at the United Nations Children’s Fund (UNICEF). Access to the data may be granted for academic research purposes upon request and with appropriate ethical approval.

